# Reliable detection of focal-onset seizures in the human anterior nucleus of the thalamus using non-linear machine learning

**DOI:** 10.1101/2020.09.18.20196857

**Authors:** Emilia Toth, Sachin S Kumar, Ganne Chaitanya, Kristen Riley, Karthi Balasubramanian, Sandipan Pati

**Affiliations:** Department of Neurology, University of Alabama at Birmingham, AL; Epilepsy and Cognitive Neurophysiology Laboratory, University of Alabama at Birmingham, AL; Centre for Computational Engineering and Networking (CEN), Amrita School of Engineering, Coimbatore, Amrita Vishwa Vidyapeetham, India; Department of Neurosurgery, University of Alabama at Birmingham, AL; Department of Electronics and Communication Engineering, Amrita School of Engineering, Coimbatore, Amrita Vishwa Vidyapeetham, India

**Keywords:** Thalamus, seizure detection, epilepsy, machine learning

## Abstract

**Objective:** There is an unmet need to develop seizure detection algorithms from brain regions outside the epileptogenic cortex. The study aimed to demonstrate the feasibility of classifying seizures and interictal states from local field potentials (LFPs) recorded from the human thalamus-a subcortical region remote to the epileptogenic cortex. We tested the hypothesis that spectral and entropy-based features extracted from LFPs recorded from the anterior nucleus of the thalamus (ANT) can distinguish its state of ictal recruitment from other interictal states (including awake, sleep).

**Approach:** Two supervised machine learning tools (random forest and the random kitchen sink) were used to evaluate the performance of spectral (discrete wavelet transform-DWT), and time-domain (multiscale entropy-MSE) features in classifying seizures from interictal states in patients undergoing stereo EEG evaluation for epilepsy surgery. Under the supervision of IRB, field potentials were recorded from the ANT in consenting adults with drug-resistant temporal lobe epilepsy. Seizures were confirmed in the ANT using line-length and visual inspection. Wilcoxon rank-sum (WRS) method was used to test the differences in spectral patterns between seizure and interictal (awake and sleep) states.

**Main Results:** 79 seizures (10 patients) and 158 segments (approx. 4 hours) of interictal stereo EEG data were analyzed. The mean seizure detection latencies with line length in the ANT varied between seizure types (range 5-34 seconds). However, the DWT and MSE in the ANT showed significant changes for all seizure types within the first 20 seconds after seizure onset. The random forest (accuracy 93.9 % and false-positive 4.6%) and the random kitchen sink (accuracy 97.3% and false-positive 1.8%) classified seizures and interictal states.

**Significance:** These results suggest that features extracted from the thalamic LFPs can be trained to detect seizures that can be used for monitoring seizure counts and for closed-loop seizure abortive interventions.

## 1. INTRODUCTION

The development of automated, reliable seizure detectors is necessary for clinical and research purposes for multiple reasons. First, an electronic seizure detector can be used to provide feedback for therapy delivery (i.e., closed-loop electrical stimulation like the responsive neurostimulator-RNS) or can trigger an alarm to notify a caregiver[1][2][3]. Second, automated detectors allow quantifying seizures and circadian patterns for an electronic seizure diary that can be used to tailor therapy[4]. Third, in research, large scale automation of seizure counts can accelerate drug development and evaluate therapy responses[5][6]. Given these translational significances, there is a continuous development of algorithms based on either machine learning or deep learning. Although the biosignal input for seizure detection has ranged from heart rate to muscle activity or scalp EEG[7][8][9], the intracranial EEG due to its higher signal to noise ratio has the highest reliability and is often used in neuromodulation therapies[10]. The developmental pipeline for seizure detection algorithms with intracranial EEG includes feature extraction from the local field potentials (LFP) recorded from the seizure onset zone, followed by training and testing. Therefore, most seizure detection algorithms are based on LFP recordings from the seizure onset zone[11]. Unfortunately, in clinical practice, the localization of seizure onset is not always possible. Treatment options like surgical resection or ablation or neuromodulation (like the RNS system) are not applicable in this cohort due to the inability to localize the seizure generation sites. Hence, mortality is higher in nonlocalizable epilepsies[12]. A preferred option in such patients has been the deep brain stimulation (DBS) which currently operates as an open loop device where stimulation parameters are pre-programmed to occur throughout the day and are not responsive to changes in the patient’s seizures resulting in various long term adverse events such as memory impairment, therapeutic ineffectiveness, etc. Thus, there is an unmet need to innovate seizure detection from brain regions outside the seizure foci or seizure onset zone. We respond to this clinical need by demonstrating the feasibility of identifying electrophysiological signatures of seizures from the human thalamus-a subcortical region that is remote to the cortical seizure onset sites.

The thalamus is not a unitary structure but a conglomeration of subnuclei, each with diverse reciprocal connectivity and functions[13]. Numerous animal studies have demonstrated that -a) recurrent limbic seizures and spikes facilitated the remodeling of the thalamocortical networks[14]; b) thalamic neuronal firings are increased with the initiation of limbic seizures[15]; and c) chemical, lesional or optogenetic deactivation of thalamic subnuclei interrupted cortical seizures[16][17][18]. For example, the anterior nucleus of the thalamus (ANT) mediates cortical-subcortical interactions between the limbic system and the brainstem[19]. These pathways are associated with a bihemispheric propagation of convulsive seizures. The thalamocortical neural activity also regulates wakefulness, sleep, consciousness, and synchronization of intracortical networks[20][21][22][23]. Prior studies have demonstrated that states of vigilance (SOV) can be distinguished using entropy or time-frequency (spectral) decomposition of EEG signals[24][25]. Based on these evidence, we tested the hypothesis that spectral and entropy-based features extracted from LFPs from the ANT can distinguish its state of ictal recruitment from other interictal states (including awake, sleep). We used two supervised machine learning approaches (random forest and the random kitchen sink) to evaluate the performance of spectral and temporal features in classifying ictal from interictal states in patients undergoing stereo EEG (SEEG) evaluation for epilepsy surgery.

## 2. METHODS

### 2.1 Study participants, inclusion criteria, and ethics

Seizures (N=79 from 10 patients) and interictal stereo EEG data (N=158 segments, i.e., 79 x2 states from sleep and awake) were obtained from patients with drug-resistant temporal lobe epilepsy (TLE) who have undergone SEEG exploration for epilepsy surgery. The demographic details of the study participants are detailed in (Table 1). The Institutional Review Board of the University of Alabama at Birmingham approved the study for recording LFPs from the thalamus during SEEG exploration (IRB-170323005). Before the surgery, informed consent detailing the thalamic implantation for research was obtained from the participants. The multi-step consenting process has been described in detail in a previous study[26]. To mitigate the risk of increased complications from an additional depth electrode placement for research purposes only, the surgeon modified the trajectory of a clinically indicated multi-contact depth electrode to sample LFPs from the insular operculum region (superficial contacts) and the ANT (deeper contacts) (Fig1A). Only one ANT was sampled per participant. The rationale for selecting the ANT was based on its -a) structural connectivity with the limbic network[19]; b) preclinical and clinical studies demonstrating recruitment of ANT in TLE[15][27]; and c) the potentials of translating the knowledge gained in the study towards the development of a closed-loop DBS therapy. Only patients with confirmed electrode localization in the ANT were included in the study

**Figure 1:**
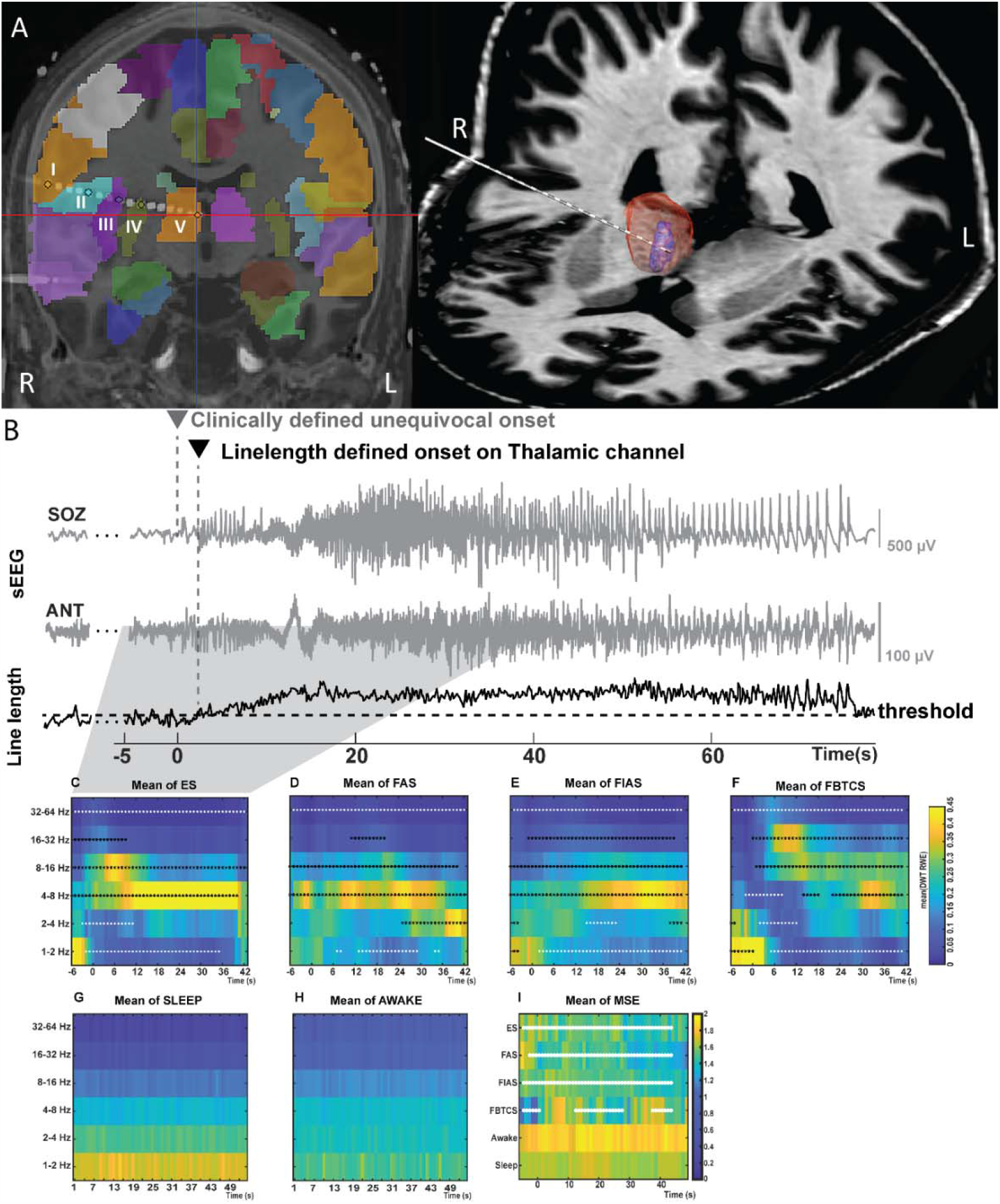
A: The trajectory of the Anterior nucleus of thalamus (ANT) implantation that followed through precentral gyrus(I), pars opercularis frontalis (II), insula (III), putamen (IV) and ANT (V). LeadDBS was used to reconstruct target location of the thalamic electrodes, while the trajectories were reconstructed and the cortical contacts were identified using iElectrodes. B: Line Length (LL) detection of seizures in the ANT. First, the unequivocal electrographic onset was identified by the clinician in the seizure onset channel (SOZ). The LL detected seizure in the ANT was subsequently confirmed with visual inspection. C, D, E, F, G, H and I: Temporal changes in discrete wavelet transform relative wavelet energy (DWT-RWE) and multiscale entropy (MSE) across first 40 seconds of seizure and interictal awake and sleep epochs in the ANT. Statistically this power spectral change during the seizure was compared with the baseline sEEG activity using Wilcoxon Rank Sum test and corrected for multiple comparisons using false discovery rate (FDR) method. C, D, E and F summarize the significant spectral power changes (increases: black asterisks and decreases: white asterisks) during various ictal states i.e., electrographic seizures (ES: z_min_<-4.1, z_max_>3.9, p’s<7.2×10^−5^), Focal Aware Seizures (FAS: z_min_<-4, z_max_>4, p’s<5.7×10^−5^), Focal Impaired Awareness Seizures (FIAS: : z_min_<-3.9, z_max_>4.2, p’s<8×10^−5^) and Focal to Bilateral Tonic Clonic seizures (FBTCS: : z_min_<-4.1, z_max_>4, p’s<5.9×10^−5^). Increase in spectral power was noted between 4-32Hz while decrease was noted majorly in 1-2Hz and 32-64Hz for all the 4 seizure types. G and H summarize the significant spectral power changes during various physiological interictal states i.e., awake and sleep. I summarizes the temporal changes in MSE during seizures and interictal period (ES: z>4.1 p’s<2.1×10^−7^, FAS: z>4.3, p’s<4.9×10^−12^, FIAS: z>4.1, p’s<2×10^−14^, FBTCS: z>4.1, p’s<4.1×10^−6^).

**Table 1:** Clinical characteristics of the study participants

| **Demographics** | **N=10** |
| --- | --- |
| • Age (years) | 42.8±13.9 |
| • Gender | M=4 |
| • Details of recording |  |
| *Number of electrodes* | 151 (R106, L45) |
| *ANT implant laterality (R: L)* | 7:3 |
| • Disease burden measures |  |
| *Age at Onset(years)* | 24.9±19.1 |
| *Duration of Epilepsy (years)* | 17.9±12.4 |
| *Frequency of focal seizures (/month)* | 12±8 (range: 2-30) |
| *FBTCS (Present: Absent)* | 8:2 |
| • MRI (Abnormal: Normal) | 5:5 |
| • PET abnormality (Abnormal: Normal) | 7:3 |
| • ANT implant laterality (R: L) | 7:3 |
| FBTCS: focal to bilateral tonic clonic seizures; PET: Positron emission tomography; ANT: anterior nucleus of thalamus |  |

### 2.2 SEEG recording

Robotic assistance (ROSA device, Medtech, Syracuse, NY) was used to plan optimal and safe trajectories for SEEG multi-contact electrode implantation (12-16 contacts per depth electrode, 2mm contact length, 0.8mm contact diameter, 1.5mm inter-contact distance, PMT^®^ Corporation, Chanhassen, MN). Intracranial video-EEG was recorded using Natus Quantum (Natus Medical Incorporated, Pleasanton, CA, sampling rate 2048Hz) (Figure 1). Signals were referenced to a common extracranial electrode placed posteriorly in the occiput near the hairline.

### 2.3 Reconstruction of depth electrodes into the brain

The pipeline and the accuracy of targeting thalamic subnuclei during SEEG have been reported previously by our group[26]. In brief, the post-implant CT volumes were co-registered to the pre-implantation MRI using affine transformation generated by FLIRT(fMRI linear image registration tool)[28]. Electrodes were localized using Lead-DBS software (www.lead-dbs.org), and trajectories were mapped using iElectrodes software[29][30]. The data was visualized using Morel’s atlas[31] (Fig1A).

### 2.4 Identification of interictal epochs and seizures in the seizure onset zone and thalamus

Seizure onset and offset in the cortex were annotated by a board-certified epileptologist (SP). Seizure onset was defined as the earliest occurrence of rhythmic or repetitive spikes in the cortex that was distinctive from the background activity, and that evolved in frequency and morphology (Fig2). The EEG onset of the seizure in the cortex was marked as “unequivocal EEG onset” (UEO). SEEG segments were clipped to include 10 minutes (min) before the UEO and 10min post-termination of the seizure. Seizures were classified into focal aware seizures (FAS), focal impaired awareness seizures (FIAS), and focal to bilateral tonic-clonic seizures (FBTCS)[32]. A seizure was classified as electrographic seizures (ES) when it lacked any clinical correlate, and the ictal EEG pattern lasted longer than 10 seconds. A minimum of three seizures selected randomly among each seizure type (ES, FAS, FIAS, FBTCS) per patient, and overall 3-10 seizures per patient were included for analyses in the study. Multiple epochs of 5-10 minutes duration of interictal SEEG were selected and labeled as “interictal state.” State of vigilance (SOV: sleep or awake) for the interictal epochs were determined based on video and visual SEEG inspection. No effort was made to classify sleep stages as the study was aimed to distinguish the thalamic ictal from interictal states. Similarly, no effort was made to determine the presence (or absence) of epileptiform spikes in the interictal epoch as the study was focused on determining the interictal state independent of presence (or absence) of a spike.

**Figure 2:**
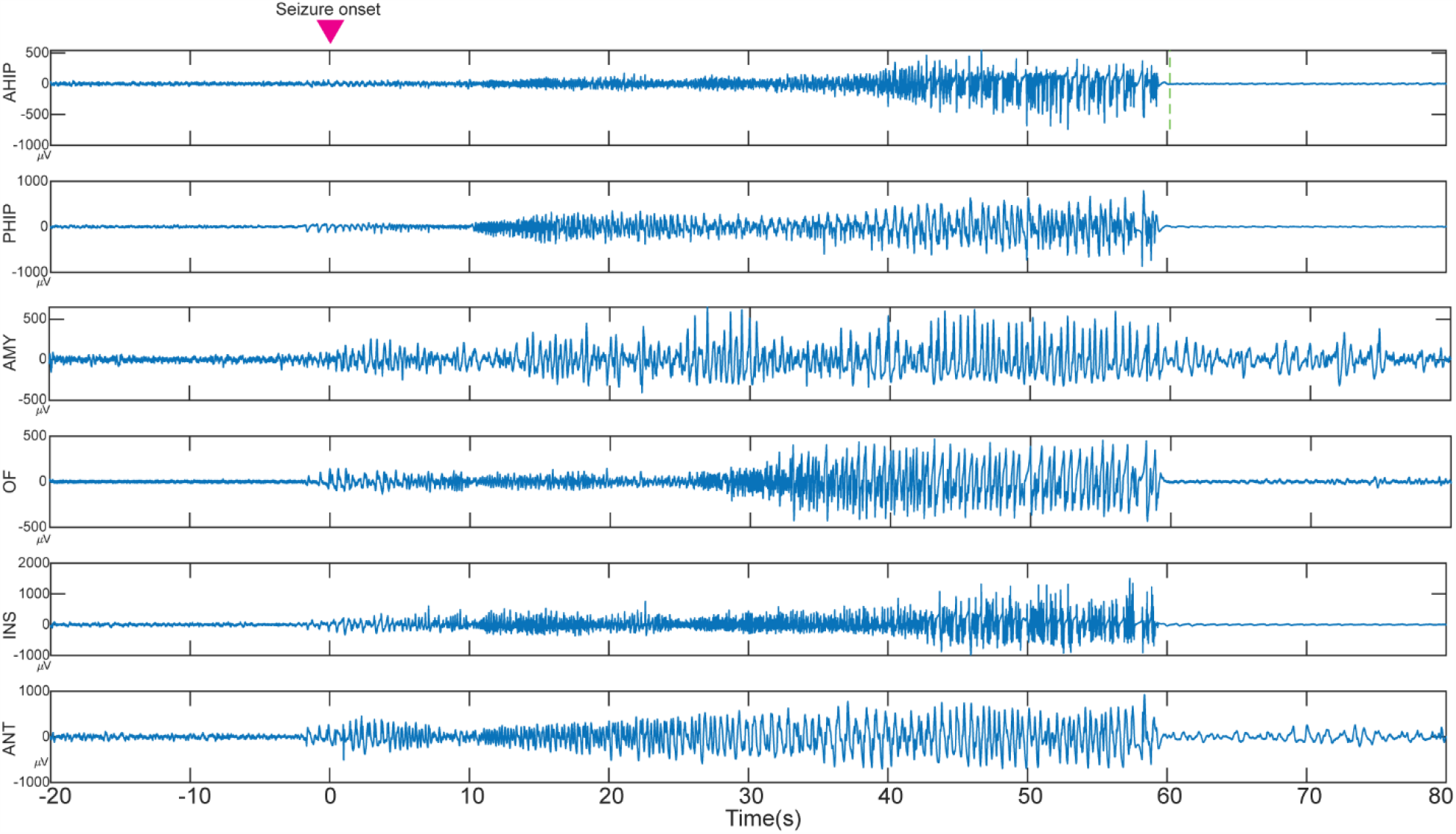
Example of a stereo EEG recording from a patient with left medial temporal lobe epilepsy. The seizure onset (time 0 sec) was characterized by low amplitude fast activity observed in the anterior and posterior hippocampus (AHIP, PHIP). At the same time rhythmic spiking was observed in the amygdala (AMY), orbitofrontal (OF) and insula (INS). Note the rhythmic spiking in the anterior nucleus of thalamus (ANT) that evolved with the progression of the seizure.

### 2.5 Seizure detection algorithm

Line length (LL) was used to detect the ictal activity objectively[33](Fig1B). The LL feature was derived as a simplification of the running fractal dimension of a signal. It measures the length of the signal in a particular window and compares it to a threshold. The length of the signal is dependent on the amplitude and frequency of the signal, making this feature highly suitable to sense changes in amplitude and/or frequency that typically occur during seizures. The change in LL during the peri-ictal period was considered significant when the measured LL in that segment was greater than 2SD compared to the mean of a 3min baseline segment for at least 10sec. Preprocessing steps involved detrending within 2s, removing DC-drift (Matlab detrend), reconfiguring into a bipolar montage by linking adjacent contacts from the proximal to distal part of the electrode, and removing 60 Hz line noise with a notch filter (2^nd^ order Butterworth zero phase shift filter –Matlab designfilt and filtfilt). Subsequently, the LL was calculated on this preprocessed data over 0.25s windows with 50% overlap as shifting windows, and the resultant LL data was smoothed using the Matlab function ‘movmean’ over 2s windows. The LL was calculated for each segment with *m* samples using the following formula:

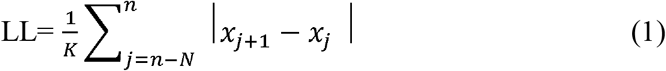

Where *n* is the number of sample points within 0.25s, and the *x*_*j*_ is the sample point.

The LL detections were merged if they were closer than 2s. When the merged detections were longer than 10s, they were considered by the detection algorithm as seizures. To improve the accuracy of the seizure detection of the entire event, on the basis of the length of the seizure, another *layer* of temporal summation was performed wherein, if the10s-merged-detections were closer than 5s, they were again merged to constitute one single seizure event. The method helped to avoid detecting them erroneously as multiple close-lying seizures, thus helping in the detection of slower ictal spiking activity.

The electrophysiological features of ANT involvement during a seizure are not well described. Thus, we have adopted a manual offline seizure detection approach based on LL features described above. LL was performed on all the cortical channels demonstrating visually determined UEO and early seizure spread in the cortex. Seizures in ANT were reported as “not detected (ND)” when the specified changes in LL as described above, were absent in the first 120s after LL detected seizure in the cortex (LL UEO).

### 2.6 Power-Spectral evaluation of the field potentials recorded from the ANT

Two methods were used to measure the ictal changes of the thalamogram, (1) calculate the power-spectra with discrete wavelet technique with relative wavelet energy (DWT-RWE) and (2) multiscale spectral entropy (MSE)[34][35][24]. The time-frequency decomposition of field potentials was performed with the DWT –‘db4’, RWE, 6 levels. The DWT provides a non-redundant, highly efficient wavelet representation and direct estimation of local energies at the different relevant scales. The motivation for selecting DWT is based on prior studies suggesting this method can be an optimal tool for online seizure detection that can be translated into the implantable neural prosthesis[11][36][37]. The DWT-RWE was calculated on 4s windows, with 3 s overlap, for 1-2 Hz, 2-4 Hz, 4-8 Hz, 8-16 Hz, 16-32 Hz, and 32-64 Hz from awake, sleep and seizure segments of the thalamic signal.

We calculated MSE to characterize the temporal predictability of a time series across several time scales in the thalamus, serving as an index of its capacity for processing information. Estimation of MSE was done in a windowed manner on short epochs of 4s each, with 75% overlap after downsampling data to 200Hz according to the original implementation of the algorithm and averaging across the 10 scales to indicate the mean MSE.

We estimated MSE for one-dimensional discrete time series (*x*) by constructing consecutive coarse-grained time series, determined by the scale factor, τ, according to equations 2 and 3,

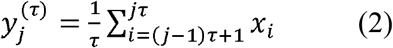

where τ is the scaling factor, 1≤j≤N/τ

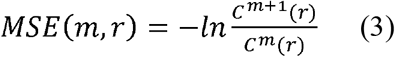

where 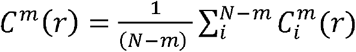, r is a coefficient of tolerance, m is the pattern length and N is the length of the data. Costa et al have shown that MSE had better statistical validity for m = 1 or 2 and r being between 0.1 and 0.25. In the present study we used m = 2, r = 0.15, 1<τ≤10.

MSE was calculated across the length of the ANT SEEG signal on moving window epochs (awake, sleep, and seizure segments) like DWT analysis.

### 2.7 Machine Learning with Random Forest(RF) and Random Kitchen Sink(RKS)

Seizures can induce changes in behavioral states (like arousal or altered vigilance) that can increase power at narrow frequency bands. To evaluate if the temporal and spectral features (MSE and DWT RWE) can distinguish the thalamic ictal state from other interictal states of vigilance (awake and sleep), we used two machine learning algorithms-RF and RKS[38][39]. Seven features, namely: DWT-RWE in 6 frequency bands and the average of the first 10 scales of the MSE of awake, sleep, and 20 seconds of seizure segments (each seizure provided 16 data points) from the ANT were used as predictive features for RF and RKS machine learning algorithms. The seizure class contained 373 data points, and the interictal class had 2850 data points (approximately 4 hours of data), each being represented by 7 features. The data was unbalanced, and hence the smaller data was split in 80%: 20% for training and testing data. 300 data points were chosen at random for training, and 73 data points were chosen for testing purposes from each class. The RF classification technique was applied using all the features to calculate the accuracy, precision (positive predictive value), and recall (sensitivity) values. To ensure that training and testing were done in an unbiased manner, the process was repeated 500 times by choosing the training and test samples randomly during each iteration, and the average values of the accuracy, precision, and recall were used. The entire procedure used 5, 10, 15, and 20 decision trees, and four sets of results were presented that indicated that we have an improved classification with an increasing number of decision trees [40]. During the construction of the trees, Gini impurity was used to make decision about which variable to split at each node. For each feature, the sum of the Gini decrease across every tree in the forest is accumulated every time that feature is chosen to split a node. The sum is divided by the number of trees in the forest to give an average called the permutation feature importance.

As a validation of the results of the RF prediction model, we tested the same features using RKS learning. The RKS method involved a nonlinear mapping of the features on to a higher dimensional space and enabled the features to become linearly separable in the higher dimension. The feature mapping was done using a real Gaussian function as the Radial basis function (RBF) kernel. This, in combination with a regularized least square algorithm for regression, allowed us to obtain a simple classifier that can be used for real-time applications. Similar to RF analysis, RKS was used to classify the given samples into two classes: seizure and interictal states (awake and sleep). RKS classification technique was applied using all the features to calculate the accuracy, precision (positive predictive value), and recall (sensitivity) values. The seven features (DWT RWE in 6 bands and the average of the first 10 scales of MSE) of each data point were transformed into a 100 dimension feature space using random feature mapping. The mapping function performed both cosine and sine transformations on each data point, thus effectively converting each data point from a seven dimension vector into a 200-dimensional vector. Using the data in the new feature space, regression analysis, and regularized least square (RLS) loss function based classification was performed using an RBF based Gaussian kernel classifier with a hold-out cross-validation. The random mapping and the classification analysis was done using Grand Unified Regularized Least Squares (GURLS)[41]. Similar to RF, the RKS was repeated 500 times by choosing the training and test samples randomly during each iteration, and the average values of the accuracy, precision, and recall were reported.

### 2.8 Statistical analysis

Since DWT-RWE showed non-normal distribution, we used the Wilcoxon rank-sum (WRS) method to test how the ictal spectral patterns differed (i.e., increase or decrease in DWT-RWE) from interictal (awake and sleep) state[42]. Five DWT RWE samples of moving windows of 4s length, with 4s overlap, were input into the WRS test to form a time-related spectral power distribution subset from the seizures with ANT recruitment separated by seizure types for every frequency band. These subset distributions were compared to awake and sleep interictal distribution with the Wilcoxon test, and False Discovery Rate (FDR) correction was applied to the resulting p values.

## 3. RESULTS

### 3.1 Safety and accuracy of thalamic implantation

Out of the 13 patients that had thalamic implants, 10 had accurate localization (77%) in the ANT and were included for subsequent analysis(Table 1). None of the patients had any ischemia or hemorrhage in the subcortical regions, as evident from the CT brain performed after the explantation of the depth electrodes. The details of the accuracy and safety of research thalamic implantation at our center have been reported elsewhere[26].

### 3.2 Seizure types and their localization

Seventy-nine seizures from 10 subjects were analyzed in this study. Based on the clinical consensus among the epileptologists, the seizure onset zone was determined to be medial temporal (2), temporal pole (1), and temporal plus (2) with the plus representing additional seizure foci (orbitofrontal or insula) outside the medial temporal region. Five patients had multifocal epilepsies with seizure foci in bilateral medial temporal regions (2), medial temporal and parietal, or insular and superior temporal gyri (2). The distribution of seizure types was: ES (26), FAS (18), FIAS (29), and FBTCS (6).

### 3.3 Line length (LL) detection latencies and spectral changes of seizures in the ANT

The detection latencies of the seizures at the seizure onset zone channels were earlier (latency: 4.57±10.28s, range: -3.97 to 63.79s) than in the ANT (latency: 16.89±21.18s, range: −0.99 to 92.98s). The mean detection latencies in the ANT varied between seizure types: FAS (34s), FIAS (12.04s), and FBTCS (5s).

Following seizure onset, the first 20 seconds in the ANT showed a progressive increase in spectral power in lower frequencies (4-16 Hz) for all seizure types (Fig1C). In addition, FIAS and FBTCS showed a sustained increase in 32-64Hz. The peak changes in theta band (4-8 Hz) were seen around 26-31 seconds after seizure onset in the cortex. Also, we observed a decrease in MSE, implying that the degree of randomness of the thalamic neural activity reduced during the time of the seizures. The changes were noted for all types of seizures (ES, FAS, FIAS, and FBTCS).

### 3.4 Electrophysiological signatures of the thalamic ictal state are distinct from interictal states

Both RF and RKS were able to parse the thalamic seizure state from interictal states. For RF, the precision (positive predictive value) and recall (sensitivity) was 93.1% and 92.8%, respectively. For RKS, the precision and recall were 96% and 94.3%, respectively. In Table 2 and Fig 3, the average values of the confusion matrices, and performance metrics that were performed over 500 iterations for both RF and RKS were highlighted. Permutation feature importance was done for the RF model in order to verify if the seven features (DWT-RWE and MSE) were relevant for the classification (Fig 3A). We observed that all the features were relevant, with the theta and beta frequency bandwidths having the highest permutation importance. For the RKS model, the features were transformed by the nonlinear RBF kernel function, and the internally transformed features were used for classification. Since a nonlinear transformation was being performed, the feature relevance analysis was non-trivial in the transformed domain and was beyond the scope of this paper. To evaluate the significant differences between the features, we used univariate ANOVA that showed that all predictors differed significantly from each other except MSE and L2-4 spectral features (Fig 3B).

**Figure 3:**
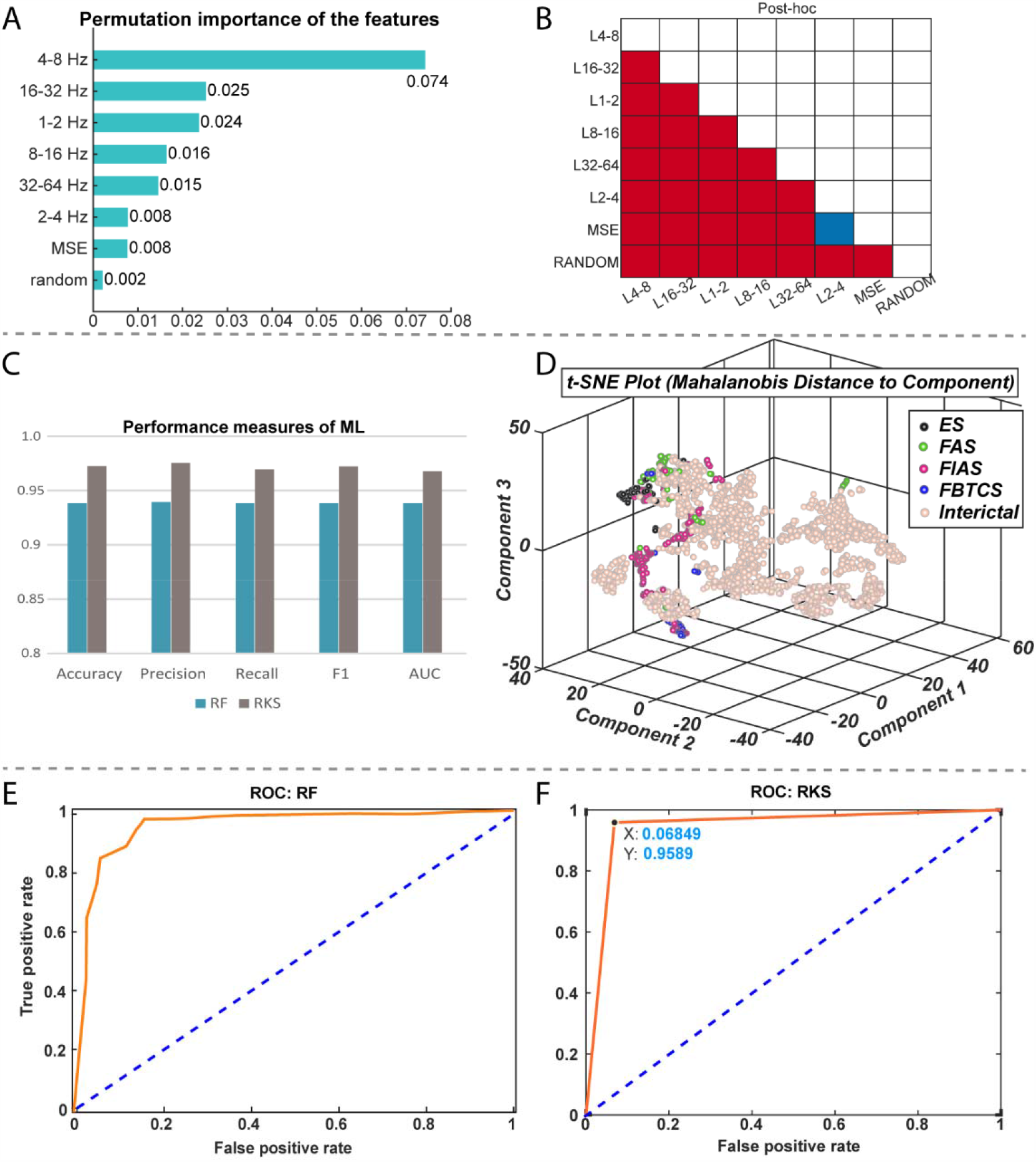
Performance measures of random forest (RF) and random kitchen sink (RKS) in classifying seizures and interictal states (awake and sleep) from electrophysiological recordings in the anterior nucleus of thalamus. A: The bar plot depicts the permutation importance of the 7 features (spectral power in 6 different frequencies and multiscale entropy (MSE) for RF classifier. Note the theta and beta frequency bandwidths had the highest permutation importance. B: Results from an univariate ANOVA that showed all the 7 features differed significantly from each other for RKS classifier except MSE and L2-4 spectral features. C: RF had an accuracy of 93.9% and a precision of 93.9%. RKS has a higher accuracy (97.3%) and precision (97.6%). D: A nonlinear dimensionality reduction using t-distributed stochastic neighbor embedding (t-SNE) showed that ictal (colored dots) spectral and MSE clusters were distinct from that of the interictal (beige dots). We also noted clustering within the ictal data and these distinct clusters (different colored dots) were because of the different seizure types (black-ES, green-FAS, red-FIAS and blue-FBTCS). Graphs E and F represent receiver operating characteristic curves (ROC) of performance of RF and RKS respectively. Both ROC curves show good separation with RF having an AUC of 0.94 and RKS with an AUC of 0.96.

**Figure 4:**
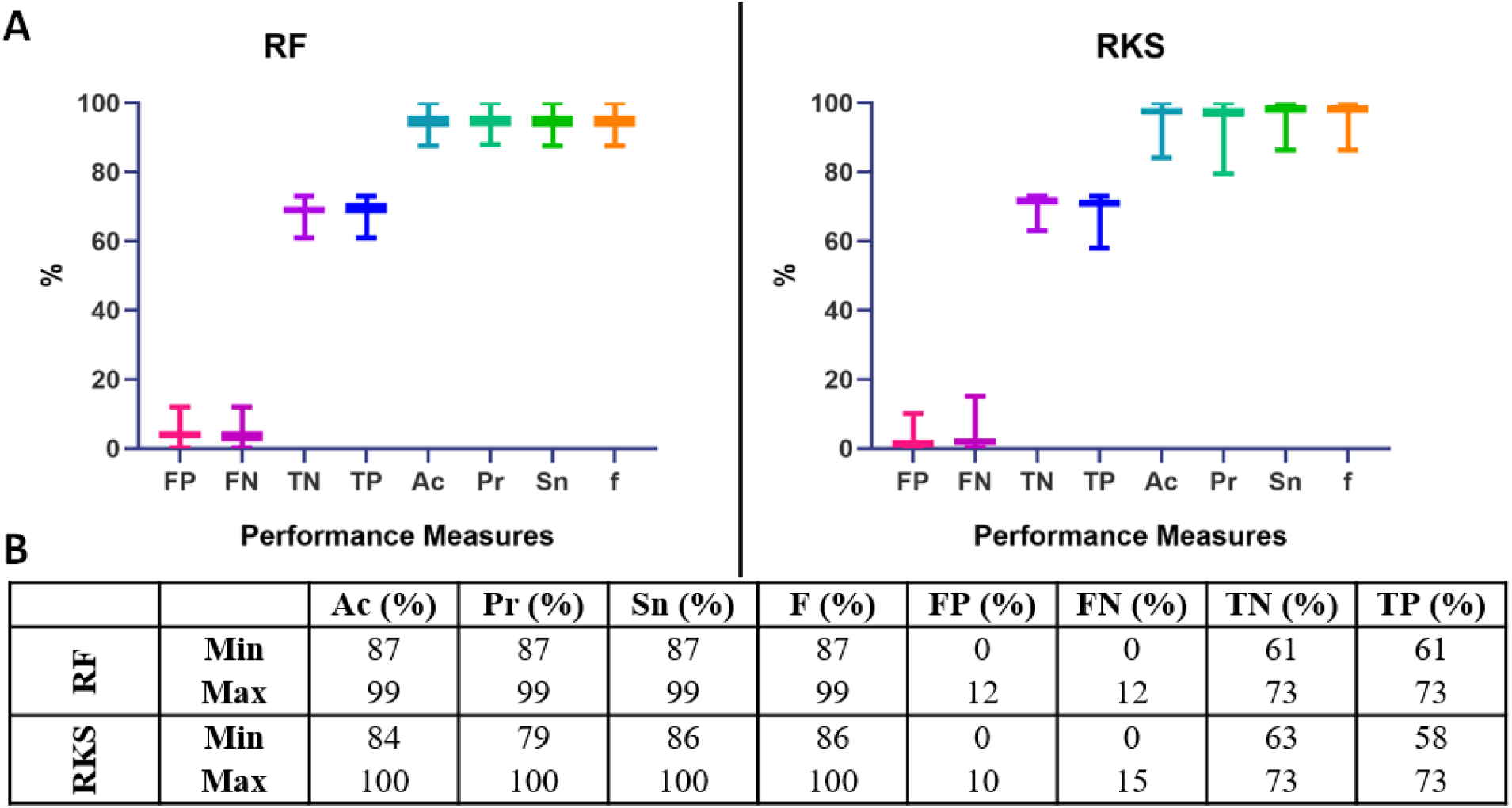
Performance (minimum and maximum) of the random forest (RF) and random kitchen sink (RKS) classifiers across the 500 iterations are enumerated. AC-accuracy; Pr-precision; Sn-sensitivity; F-f measure; FP-false positive; FN-false negative; TN-true negative; TP-true positive.

After establishing the relevance of the seven features in classifying seizures and the interictal data, the seven-dimensional data was visualized as a two-dimensional map using t-distributed stochastic neighbor embedding (t-SNE) algorithm. The 2-D map of the features was plotted using t-SNE with Mahalanobis distance metric (Fig 3D). It can be observed that the DWT RWE and MSE clusters during seizures were distinct from the interictal DWT RWE and MSE, thus strengthening the probability of correct classification of thalamic states as seizure or inter-ictal.

Overall, these results demonstrate that features extracted in the time and spectral domain from the thalamic LFPs can be used to classify seizures and interictal states in focal epilepsy.

## 5 DISCUSSION

For the past two decades, there has been a significant increase in the efforts at developing seizure detection classifiers, nearly all of which were recordings from the cortical brain structures. However not always is a clear cortical focus evident, as in the case of cryptogenic epilepsies. There is a paradigm shift in understanding epilepsy as a focal disorder to one of a network disorders. Hence, targeting key structural substrates in such network disorders can help reduce the risk of having a seizure or prevent the seizure from spreading to other parts of the brain. This becomes increasingly relevant in drug resistant MRI negative focal epilepsies. Hence it is pertinent to use human electrophysiological signals from subcortical structures to detect seizures states, particularly the human thalamus. Current thalamic DBS therapy administers continuous stimulation and has only as recently as in 2017 been approved by the U.S. Food and Drug Administration (FDA) as an add-on therapy for epilepsy. By this time, the closed-loop neuromodulation RNS system was already in clinical use for 5 years (approved in 2012). RNS unlike DBS, delivers stimulation when it detects the beginnings of seizure like activity but this is limited to the cortical level. DBS suffers the limitation of being an open loop stimulation given either continuously or in cycles. Preliminary attempts at closing this loop have shown promising results, with patients not reporting the untowardly adverse effects on mood, memory or behavior as reported in long-term DBS therapy. However, for effective detection of seizures in the thalamus, the brain computer interface as the one in the RNS detection system, should rely on more accurate detection algorithms. Hence in this study we have demonstrated how machine learning classifiers can segregate frequency specific power spectral changes in the thalamus occurring during early seizure from baseline interictal states. Machine learning algorithms have been at the forefront as they provided higher accuracy in detecting seizures while minimizing the false positive rate. Using supervised machine learning tools, we demonstrated the feasibility of classifying seizures and interictal states from LFP recordings from the ANT. The performance metrics for the learning algorithms were comparable to the existing seizure classifiers[11][43], although the notable finding in our study was the demonstration of seizure detection outside the seizure onset region. Currently, detection algorithms from non-brain signals continue to have a poor sensitivity ranging between 57-64% for wearable electrocardiography (ECG) and 33% for wearable photoplethysmography sensors monitoring pulse rate variability[44][45]. Among the brain-signals, multi-channel scalp EEG data classifiers have performed better with an average accuracy of 99.32%, the sensitivity of 99.41%, and specificity of 95.25%[10][9]. Most scalp EEG based detection studies had sensitivity between 83%-96%, accuracy between 84%-100%, and false detection rate of as low as 0.15-0.17 events/hour[11][10]. Most invasive EEG studies implementing classifiers to detect seizures from the seizure onset zone had sensitivity between 86%-100%, accuracy between 70% and 100%, and false detection rate of as low as 0 events/hour[11][43]. Comparable to these, our thalamic based detections performed relatively well in detecting seizures among both the random forest (sensitivity 92.8%, accuracy 92% and false-positive 7.03) and random kitchen sink classifiers (sensitivity 94.3%, accuracy 95% and false-positive 4.14).

### 5.2 The emergence of theta rhythm in the ANT during seizures

Although multiple spectral changes characterized the thalamic ictal state, the emergence of theta band was significantly prominent and sustained for seizures that remained focal. Our findings are in agreement with recent studies that demonstrated increased power in lower frequencies in the ANT at the seizure onset[46]. Vertes et al. demonstrated the presence of rhythmic burst firing of neurons in the ANT that was synchronous with hippocampal theta rhythm, and the rhythm resonated within the Papez circuit (including medial septum hippocampus and ANT)[47]. Prior studies have demonstrated that theta rhythm is associated with increased seizure threshold, is anti ictogenic, and synchronization in theta rhythm can ameliorate epileptic discharges[48][49]. Further work is necessary to establish the role of theta rhythm in seizure propagation.

### 5.3 Clinical implication of developing thalamic seizure detector

With the increase in multifocal and, difficult-to localize epilepsies, there is a need to develop effective neuromodulation therapies as surgical resection is often unavailable. The ANT mediates cortical-subcortical interactions between the limbic system and the brainstem. These pathways are associated with a generalization or bihemispheric propagation of convulsive seizures[50]. Thus the thalamic hub is an attractive target for modulating the distributed epileptogenic network[26][51]. Indeed stimulation of the ANT confirmed modulation of the limbic system. Furthermore, recent case studies highlighted the feasibility of the RNS system in modulating the thalamocortical nodes during seizures[52][53]. Our work is a step forward in identifying reliable seizure markers in the thalamus.

### 5.4 Limitations of machine learning model

In the present study, the data was trained on seizure clippings as discrete events, and the event detection was considered as a classifier and not as a prediction. Future studies should validate the findings on an extended period of EEG recordings from the thalamic subnuclei. Secondly, the features used for machine learning were limited to entropy and spectral measures. Other features like the synchronization patterns between the seizure onset zone and the thalamus are likely to influence the detection rate of seizures. Lastly, we noted that these machine learning tools were time expensive, where our data sets were trained and tested over hours. For neuromodulation based on online event detection, the algorithms need to perform faster while retaining the higher accuracy and sensitivity.

## 6. CONCLUSION

For closed-loop neuromodulation therapies, seizure detection algorithms are often developed by extracting features from the LFPs recorded from the seizure onset zone. In the present proof-of-concept study, we demonstrated that focal onset seizures could be detected in the thalamus-a subcortical structure that is outside the epileptogenic cortex. Using two supervised machine learning algorithms, we demonstrated the feasibility of classifying seizures from other interictal states of vigilance (awake, sleep) with higher accuracy (RF=92%; RKS= 95%) and sensitivity (RF=92.8%; RKS=94.3%). Overall, these results suggest that field potentials recorded from ANT can be potential biomarkers for classifying states that can be targeted for intervention.

## Data Availability

Provided at reasonable request after final publicaiton of article.

## Abbreviations

ASD: Anti-seizure drugs
DWT: Discrete Wavelet Transfer
EI: Epileptogenecity Index
ES: Electrographic Seizures
FAS: Focal Onset Aware Seizures
FBTCS: Focal to bilateral tonic-clonic seizures
FIAS: Focal onset seizures with impaired awareness
LL: Line Length
MSE: Multiscale Entropy
RF: Random Forest
RKS: Random Kitchen Sink
RWE: Relative Wavelet Energy
SOZ: seizure onset zone
SEEG: stereo-electroencephalography
TLE: Temporal lobe epilepsy
UEO: Unequivocal electrographic onset

## ACKNOWLEDGMENTS

ET, GC, and SP would like to acknowledge the continuous support from the USA National Science Foundation Grant (NSF RII-2 FEC OIA-1632891). SP and GC would also like to acknowledge support from NIH (1RF1MH117155-01). We would like to thank Diana Pizarro and Auriana Irannejad for helping in data organization.

